# Risk Factors and Risk Score (NOAM score) for Peripartum Cardiomyopathy

**DOI:** 10.64898/2026.02.05.26345708

**Authors:** Hezzy Shmueli, Talish Razi, Tal Biron-Shental, Yarden Cohen, Avraham Shotan, Doron Zahger, Ronen Arbel, Talia Lanxner Battat, Shimrit Yaniv-Salem, Uri Elkayam, Dan Yamin

## Abstract

**Introduction:** Peripartum cardiomyopathy (PPCM) is a life-threatening pregnancy-associated cardiomyopathy characterized by left ventricular systolic dysfunction. Due to its relative rarity, risk factors are not well established, and a validated risk score is lacking.

**Methods:** We conducted a retrospective cohort study, analyzing 317,892 deliveries between January 2014-December 2024. PPCM cases were identified by individual clinical chart review and echocardiographic validation. Multivariate logistic regression identified independent predictors. A post hoc stratified analysis created five risk categories to estimate PPCM incidence.

**Results:** We identified 107 PPCM cases (1:2,970 deliveries). Significant risk factors included cardiovascular disease or arrhythmias, odds ratio (OR) 8.18 (95% confidence interval [CI], 4.34 to 14.2); preeclampsia, OR 7.50 (95% CI, 4.69 to 11.7); Women of African ancestry, OR 4.39 (95% CI, 2.32 to 7.64); first delivery, OR 3.38 (95% CI, 2.25 to 5.08); multifetal pregnancy, OR 2.76 (95% CI, 1.36 to 5.07); and maternal age older than 35 years, OR 2.30 (95% CI, 1.50 to 3.48). In the post hoc stratified analysis, risk groups demonstrated marked heterogeneity, with risk ranging from 1 per 25,000–7,692 deliveries in the lowest risk group to 1 per 163–65 deliveries in the highest risk group.

**Conclusion:** We identified risk factors for PPCM and developed the NOAM Score, a simple and clinically applicable tool that integrates several domains—preeclampsia, cardiovascular disease, hypertension, Women of African ancestry, primiparity, advanced maternal age and multifetal pregnancy. This framework stratifies women across a wide spectrum of risk and may support earlier identification, risk-guided monitoring, and targeted preventative care.

## Introduction

Peripartum cardiomyopathy (PPCM) is a pregnancy-associated cardiomyopathy that represents a significant cause of non-obstetric morbidity and mortality (1, 2). Thus, early recognition and diagnosis of PPCM is critical. PPCM typically presents toward the end of pregnancy or in the months following delivery, manifesting as heart failure secondary to left ventricular (LV) systolic dysfunction. At the time of diagnosis and for most of the affected women, the average Left ventricular ejection fraction (LVEF) is around 30% (3). PPCM prevalence varies significantly by geography – ranging from as high as 1 in 100 pregnancies in Nigeria to as low as 1 in 20,000 in Japan. (4, 5)

Recognizing and diagnosing PPCM is critical, as its consequences can be devastating for both mother and fetus (2). However, symptoms such as effort intolerance and dyspnea can be easily misattributed to common benign third trimester or early post-partum complaints, leading to misdiagnosis and delay in appropriate care(1).

These considerations are pertinent not only for the index pregnancy and postpartum period, but also for subsequent pregnancy (SSP) and family planning decisions as well as the need for long term follow up.(6–8)

The existing literature largely focuses on prognostic factors after diagnosis of PPCM, particularly the likelihood of recovery and the risk associated with SSP. There is previous data on suggested risk factors for PPCM, but no studies to date have attempted to assemble a risk prediction model / score for prediction of future PPCM.

The aim of this study was, therefore, to identify risk factors associated with the development of PPCM, using a large and heterogeneous obstetric cohort to create a population-based predictive screening model. This model would help to identify and classify high risk women that will benefit from intensive follow up and treatment, and if addressed early, might reduce the incidence of PPCM.

## Methods

We conducted a retrospective cohort study using data from "Clalit Health Services" (CHS), Israel’s largest healthcare provider, covering approximately 50.7% of the population. The dataset included deliveries from January 2014 to December 2024 across CHS-affiliated hospitals nationwide, representing a heterogeneous population.

Clinical and demographic data were extracted from CHS’s centralized electronic health records, including diagnostic codes (based on the International Classification of Diseases, 10th Revision, Clinical Modification [ICD-10-CM]), medical history, treatments, surgical procedures, hospitalizations, and mortality. The study was approved by the CHS Community Helsinki Committee and the CHS Data Utilization Committee, and was exempt from the requirement for informed consent.

We identified all of the deliveries during the study period using diagnosis and procedural codes. Women with several deliveries were included until an event occurred. To detect PPCM cases, we screened for relevant codes from third trimester antepartum to 6 months postpartum, including PPCM-specific codes as well as codes related to acute HF and new-onset cardiomyopathy, in order to capture miscoded cases.

We excluded women with known heart failure (HF) and / or cardiomyopathy and HF-related codes in the year preceding delivery. Data collection began one year prior to pregnancy to distinguish preexisting cardiomyopathy from peripartum cases. All suspected PPCM cases were reviewed independently by a senior cardiologist (HS) and a senior obstetrician - gynecologist (TBS), who evaluated each clinical record and echocardiogram to confirm diagnosis and exclude alternative etiologies.

We pre-specified candidate covariates based on clinician experience and prior evidence. Advanced maternal age, African ancestry, hypertensive disorders of pregnancy (including pre-eclampsia and gestational hypertension), multifetal gestation, obesity, diabetes, smoking, and parity-related factors have all been reported in association with PPCM or maternal cardiovascular risk (9–11). We included sector (General Jewish, Ultra-Orthodox, Arabic) and socioeconomic score to capture social determinants, given reported links between socioeconomic disadvantage and worse maternal cardiovascular outcomes (12, 13). Low-dose aspirin use was retained as a proxy for high-risk obstetric management (per guideline indications for pre-eclampsia prevention) (14, 15).

Endometriosis was included for adjustment/exploration due to its emerging association with long-term cardiovascular disease risk, although its specific relationship to PPCM remains uncertain. Advanced maternal age, particularly 35 years and above, has been consistently identified as a high risk pregnancy (9).

### Statistical Analysis

We first conducted a univariate analysis between participants with PPCM and those without. For each prespecified covariate, we calculated the odds ratio (OR) for PPCM with its 95% confidence interval (CI) and p-value. Because PPCM occurs within a narrow late-pregnancy to early-postpartum window, event timing shows little variability; therefore, we used logistic regression rather than a time-to-event model. Hence, we constructed a multivariable logistic regression model to identify independent predictors of PPCM. Given the rarity of PPCM and to avoid overfitting, only variables with a p-value < 0.05 in the univariate analysis were included in the model, to limit overfitting given the rarity of PPCM. Model performance was assessed using standard diagnostic measures, and multi-collinearity was evaluated to ensure stability of the estimates. All analyses were conducted using R, version 4.5.0.

To support clinical interpretation, we conducted a post hoc analysis to stratify the population into PPCM risk categories. Given the relatively small number of PPCM cases in the cohort, we applied a stringent significance threshold of p < 0.005 in the prior analysis to minimize the risk of false-positive findings. The six variables meeting this criterion formed the basis for the stratification.

Each pregnancy was classified into a risk category based on the presence or absence of six key risk factors (age ≥35 years, first delivery, women of African ancestry, multifetal pregnancy, preeclampsia, and cardiovascular disease/arrhythmias). Women with none of these risk factors formed the very-low-risk group. The remaining pregnancies were grouped hierarchically according to observed incidence of PPCM, and adjacent groups were combined until each category contained at least 10,000 women and 10 or more PPCM cases. This approach yielded five mutually exclusive categories (very low, low, elevated, serious, and critical). For each category, we estimated the incidence of PPCM per 1,000 pregnancies and calculated 95% confidence intervals (CIs).

## Results

A total of 318,746 women delivered during the study period, of whom 652 were excluded for prior diagnosis of HF and / or cardiomyopathy or age < 18. Of 317,892 eligible deliveries, 202 were initially identified as PPCM cases. Following adjudication for each case by two senior physicians (HS, TBS), 95 cases were excluded mostly due to misdiagnosis (no LVEF impairment on echocardiography, previous documented cardiomyopathy that was not coded properly etc.), leaving 107 confirmed PPCM cases (1 in 2970 deliveries). The control group included 317,785 women (Figure 1).

**Figure 1.**
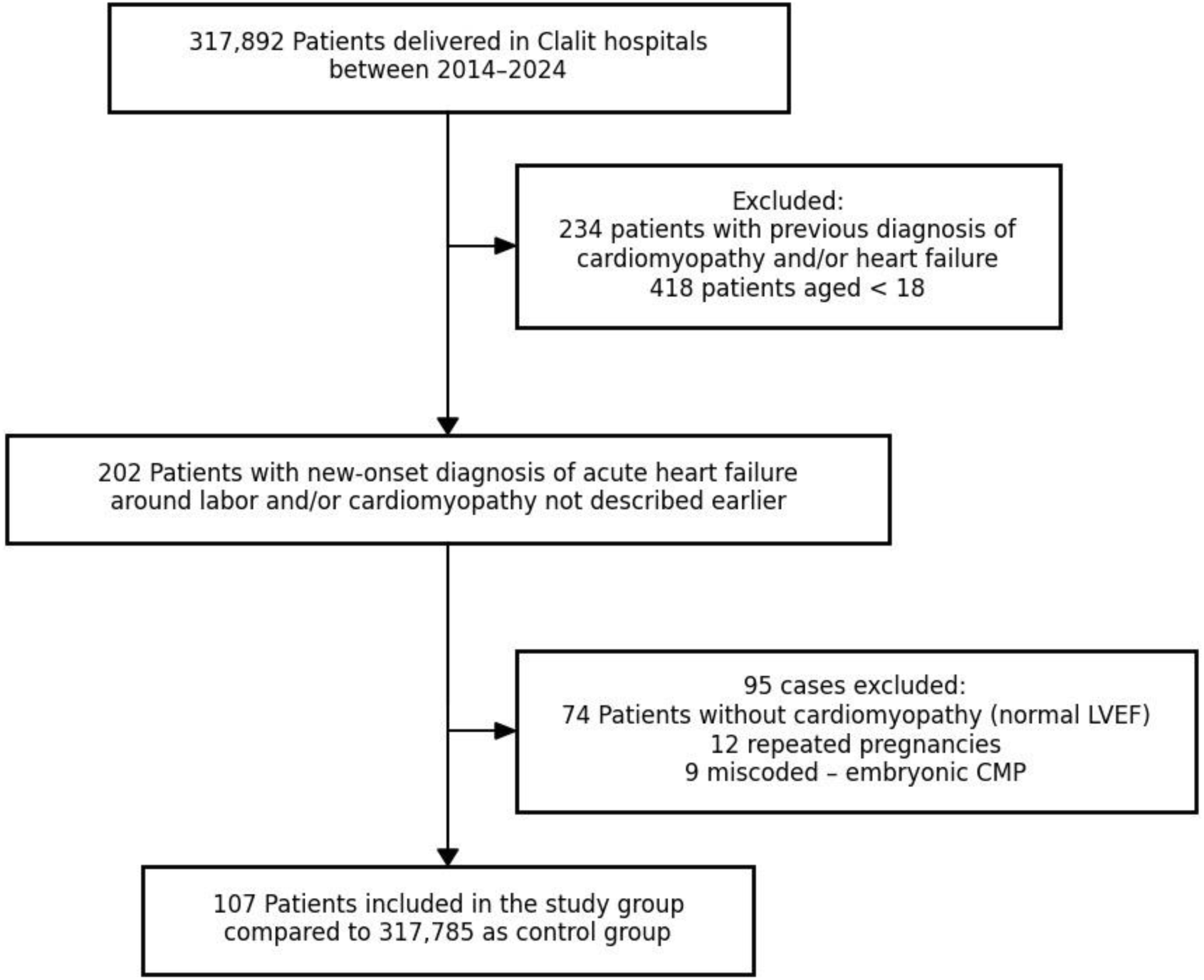
Flowchart of patient selection process for the study.

The characteristics of the study group compared to the control group are detailed in Table 1. PPCM patients were older, more likely to be of Women of African ancestry, and had higher rates of diabetes mellitus, obesity, hypertension, and aspirin use. PPCM cases more frequently had a first delivery (OR 2.93, 95% CI 2.00–4.30), experienced pre-eclampsia (OR 13.2, 95% CI 8.46–19.9) and had a history of prior caesarean section (OR 1.50, 95% CI 0.91–2.37).

**Table 1.** Baseline Characteristics of the Study Population According to PPCM Status.

|  | Without PPCM<br>N = 317,785 | With PPCM<br>N = 107 | Odds Ratio<br>(95% Confidence Interval) | p.value |
| --- | --- | --- | --- | --- |
| **Age > 35, n (%)** | 77,830 (24%) | 43 (40%) | 2.07 (1.40, 3.04) | **<0.001** |
| **Sector, n (%)** |  |  |  |  |
| General Jewish | 195,314 (61%) | 73 (68%) |  |  |
| Arabic | 107,409 (34%) | 32 (30%) | 0.8 (0.52, 1.20) | 0.3 |
| Ultra-orthodox | 15,062 (4.7%) | 2 (1.9%) | 0.36 (0.06, 1.13) | 0.15 |
| **Socioeconomic score, Median (IQR)** | 5 (3, 7) | 5 (3, 7) | 0.98 (0.90, 1.06) | 0.6 |
| (Missing) | 36,301 | 16 |  |  |
| **Women of Ethiopian origin, n (%)** | 9,422 (3.0%) | 13 (12%) | 4.53 (2.42, 7.79) | **<0.001** |
| **Malignancy, n (%)** | 4,085 (1.3%) | 2 (1.9%) | 1.46 (0.24, 4.61) | 0.6 |
| **Diabetes Mellitus, n (%)** | 2,433 (0.8%) | 5 (4.7%) | 6.35 (2.24, 14.1) | **<0.001** |
| **Obesity, n (%)** | 31,507 (9.9%) | 17 (16%) | 1.72 (0.99, 2.80) | **0.041** |
| **Smoking status, n (%)** |  |  |  |  |
| No smoker | 257,071 (81%) | 89 (83%) | — |  |
| Former Smoker | 26,065 (8.2%) | 12 (11%) | 1.33 (0.69, 2.33) | 0.4 |
| Current Smoker | 34,649 (11%) | 6 (5.6%) | 0.5<br>(0.19, 1.05) | 0.1 |
| **CVD and Arrhythmias, n (%)** | 5,168 (1.6%) | 13 (12%) | 8.37 (4.46, 14.4) | **<0.001** |
| **Hypertension, n (%)** | 1,570 (0.5%) | 6 (5.6%) | 12 (4.67, 25.0) | **<0.001** |
| **Endometriosis, n (%)** | 2,341 (0.7%) | 2 (1.9%) | 2.57 (0.42, 8.09) | 0.2 |
| **Gestational hypertension, n (%)** | 4,272 (1.3%) | 3 (2.8%) | 2.12 (0.52, 5.62) | 0.2 |
| **Gestational diabetes, n (%)** | 11,351 (3.6%) | 7 (6.5%) | 1.89 (0.80, 3.78) | 0.1 |
| **Aspirin, n (%)** | 7,212 (2.3%) | 10 (9.3%) | 4.44 (2.17, 8.09) | **<0.001** |
| **First delivery, n (%)** | 91,455 (29%) | 58 (54%) | 2.93 (2.00, 4.30) | **<0.001** |
| **Previous cesarean section, n (%)** | 44,401 (14%) | 21 (20%) | 1.5 (0.91, 2.37) | 0.094 |
| **Multifetal pregnancy, n (%)** | 7,621 (2.4%) | 11 (10%) | 4.66 (2.36, 8.32) | **<0.001** |
| **Pre-eclampsia, n (%)** | 8,723 (2.7%) | 29 (27%) | 13.2 (8.46, 19.9) | **<0.001** |

\* **PPCM** - peri partum cardiomyopathy
\*\* **CVD** - cardiovascular disease

The multivariable analysis was based on the significant parameters found in the univariate analysis and is shown in Table 2. The main independent predictors of PPCM included sociodemographic factors such as maternal age >35 years (OR 2.27, p<0.001) and Women of African ancestry (OR 4.51, p<0.001) and pre-existing conditions including CVD – mainly mitral valve regurgitation and premature atrial and ventricular ectopy (OR 8.11, p<0.001) and pre-eclampsia (OR 7.2, p<0.001).

**Table 2.** Multivariate Logistic Regression Analysis for Predictors of Peripartum Cardiomyopathy.

|  | **OR<sup>1</sup>** | **95% CI<sup>1</sup>** | **p-value** |
| --- | --- | --- | --- |
| **Age > 35** | 2.3 | 1.50, 3.48 | **<0.001** |
| **Women of Ethiopian origin** | 4.39 | 2.32, 7.64 | **<0.001** |
| **Diabetes Mellitus** | 2.5 | 0.84, 6.00 | 0.063 |
| **Obesity** | 1.23 | 0.69, 2.08 | 0.5 |
| **CVD and Arrhythmias** | 8.18 | 4.34, 14.2 | **<0.001** |
| **Hypertension** | 3.18 | 1.16, 7.33 | **0.013** |
| **Aspirin** | 1.51 | 0.70, 2.97 | 0.3 |
| **First delivery** | 3.38 | 2.25, 5.08 | **<0.001** |
| **Multifetal pregnancy** | 2.76 | 1.36, 5.07 | **0.002** |
| **Pre-eclampsia** | 7.5 | 4.69, 11.7 | **<0.001** |
| <sup>1</sup> OR = Odds Ratio, CI = Confidence Interval, CVD – cardiovascular disease |  |  |  |

In accordance with the model, we also developed a simple risk score to guide clinicians in estimating PPCM risk. No listed factors were assigned 0 points; age ≥35 years or first delivery were assigned 1 point; Women of African ancestry or multifetal pregnancy 2 points each; and eclampsia or CVD or arrhythmias 7 points each, as another 1-point risk factor does not change the category, but all other combination enhances the risk. Based on these clinically established risk factors, we stratified the cohort into five mutually exclusive categories (Table 3). Risk groups demonstrated marked heterogeneity, with risk ranging from 1 per 25,000–7,692 deliveries in the lowest-risk group (score = 0) to 1 per 163–65 deliveries in the highest-risk group (score ≥8). More specifically, the critical-risk group (preeclampsia or cardiovascular disease/arrhythmias combined with either age ≥35 years and first delivery, women of African ancestry, or multifetal pregnancy; score >8) comprised 0.6% of the population and had a PPCM risk of 9.43 per 1,000 deliveries (95% CI, 6.15 to 15.37). The serious-risk group (preeclampsia or cardiovascular disease/arrhythmias alone; score = 7-8) represented 3.7% of the population, with a risk of 2.05 per 1,000 deliveries (95% CI, 1.23 to 2.94). The very-low-risk group (no listed risk factors; score = 0) comprised 45.8% of the population and had the lowest observed risk, 0.07 per 1,000 deliveries (95% CI, 0.04 to 0.13), as shown in Figure 2.

**Figure 2.**
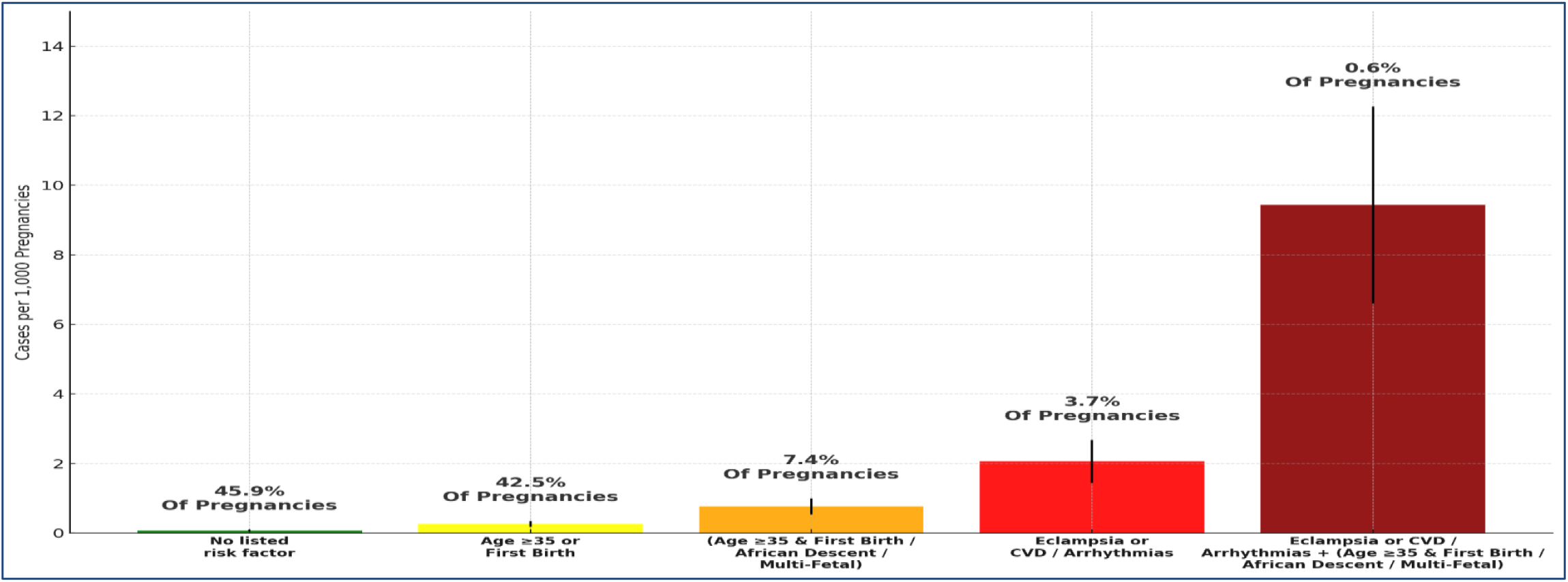
Events per 1,000 Pregnancies and Group Size Across Stratified Risk Categorie.

**Table 3.** Events per 1,000 Pregnancies and Group Size Across Stratified Risk Categorie.

| Risk Stratification by Group | Risk Score | N | % Of Total Population | Risk | Events | Risk per 1000 | Risk CI 95% per 1000 |
| --- | --- | --- | --- | --- | --- | --- | --- |
| No listed risk factor | 0 | 145,869 | 45.8% | Minimal | 11 | 0.07 | 0.04 - 0.13 |
| Age ≥35 / First delivery (1p. each) | 1 | 135,000 | 42.5% | Low | 36 | 0.27 | 0.19 - 0.37 |
| (Age ≥35 & First delivery) / Women of Ethiopian origin / Multi-Fetal (2p. each) | 2-6 | 23,497 | 7.4% | Elevated | 18 | 0.76 | 0.42 - 1.12 |
| Eclampsia / CVD / Arrhythmias (7p. each) | 7-8 | 11,672 | 3.7% | Serious | 24 | 2.05 | 1.23 - 2.94 |
| Eclampsia or CVD or Arrhythmias + (Age ≥35 & First delivery / African Descent / Multi-Fetal) | >8 | 1,908 | 0.6% | Critical | 18 | 9.43 | 6.15 - 15.37 |
| **Total** | - | **317,892** | **100%** |  | **107** |  |  |

\* CVD - cardiovascular disease, 1p. = 1 point, 2p = 2 points, 7p = 7 points

## Discussion

In this large, population-based cohort of 317,892 deliveries over 10 years, we identified major demographic and clinical risk factors for peripartum cardiomyopathy and developed a risk score that stratifies women across a broad spectrum of risk. The NOAM (**N**umeric cardio-**O**bstetric **A**ssessment for **M**aternal PPCM) distinguishes risk ranging from very-low-risk pregnancies, with a risk as low as 0.07 per 1,000 deliveries, up to critical-risk pregnancies, with a risk approaching 9.4 per 1,000 deliveries. These findings highlight both the relative rarity of PPCM in the general population and the existence of clearly defined subgroups with substantially higher risk, underscoring the potential value of targeted surveillance and early intervention in obstetric and cardiology practice.

We found that the most powerful risk factors for PPCM were background CVD (which referred mostly to mild and moderate mitral valve regurgitation and premature atrial and ventricular ectopic beats), Women of African ancestry, preeclampsia, primiparity, hypertension and advanced maternal age. These findings align partially with previous reports, such as by Lee et al., who reported some similar findings in South Korea (5); however in our current study we discovered novel risk factors. Furthermore, we took the descriptive statistics to the next step and adjusted the findings into a clinical diagnostic tool, for prediction of the risk to develop PPCM and attempt to modify it.

Some of the risk factors were previously described in different populations, but unique to our population we found a higher risk of developing PPCM in Jews of Ethiopian descent similar to that described in black African and African American women. In many African countries, PPCM is a leading cause of morbidity and HF hospitalizations. In Nigeria there are specific regions with incidence of 1:96 pregnancies, but it’s on the rise also in South Africa, Tanzania and Uganda. However, evidence on incidence and outcomes specifically from Ethiopia is lacking (16). This highlights, especially in comparison to Bedouin population in southern Israel, that the risk probably reflects genetic predisposition and does not correlate to low socioeconomic status, which these two populations share in Israel on the one hand, but the Bedouin origin was not found to be significant risk. A different profile of PPCM was previously described by Irizarry et al. in African American vs non-African American women in 2017 (17). They reported 220 African American women who were diagnosed with PPCM and compared their characteristics to non-African ancestry women. They showed the African American women had the diagnosis at a younger age (27.6 vs 31.7 years, P < 0.001), were diagnosed later in the postpartum period, and were more likely to present with a LVEF less than 30% compared with non-African American women (48 [56.5%] vs 30 [39.5%], P = 0.03). African American women’s condition was also more likely to worsen after the initial diagnosis, and they were twice as likely to fail to recover or to recover much more slowly despite adequate treatment.

Two of the most powerful predictors in our study were prior CVD as defined above and preeclampsia. These findings may correlate with the suggestion that PPCM and gestational hypertension / eclampsia lay on the same etiologic spectrum of endothelial dysfunction (and probably can explain the significant use of aspirin in these groups as seen in Table 1). Both conditions feature impaired vasodilation (caused by increased vascular sensitivity to vasoconstrictors and reduced production of vasodilators like nitric oxide and prostacyclin), increased vascular tone, inflammation, and a pro-thrombotic State (18–21). While preeclampsia is triggered by pregnancy-specific placental factors, CVD is driven by chronic lifestyle and metabolic risk factors(22). As for other “traditional” cardiovascular risk factors, some of them did correlate with high OR for PPCM such as hypertension. At the same time, smoking did not emerge as a PPCM risk factor, in agreement with a previously described association between smoking and a lower risk of preeclampsia (23, 24).

A few previous studies tried to predict the risk of developing PPCM. Lewey et al. described a cohort of 220 patients with hypertensive disorders of pregnancy who developed PPCM early postpartum. They concluded that early diagnosis was associated with a better outcome (25). Goli et al. discussed the genetic and phenotypic landscape of PPCM and demonstrated that predisposition to heart failure from TTN genes is an important risk factor for PPCM (26). However, these genetic data are not commonly available as a clinical screening tool, particularly not in developing countries. Davis et al. described a delivery-time prediction model to develop PPCM with associated risk factors such as Diabetes mellitus, obesity, mood disorders, and lower socioeconomic status (27). These risk factors did correlate with some of our findings, however their prediction relied on women presented at the time of delivery, whereas our prediction model is directed to early diagnosis of at-risk population in order to try and prevent the acute HF presentation at delivery. Interestingly, two years ago Jung et al. described a 12-lead electrocardiogram-based deep learning model to screen for PPCM. This tool managed to identify 6% of 204 patients, leading to 1-lead noninvasive and effective algorithm of identifying cardiomyopathies occurring during the peripartum period(28). However, this issue seems to need more validation as a very recent study by Kinaszczuk et al. They described a two-way artificial intelligence (AI) tool (ECG and AI-stethoscope) that managed to achieve area under the curve of 0.94 and 0.98 respectively in identifying LV systolic dysfunction among 100 women at reproductive age, validated by echocardiography. However, this model was not related to PPCM (29).

## Limitations

This is a retrospective clinical data study, which relies on coding. Hence, theoretically the disease can be underdiagnosed, surely in the first years of the study, because as the years progressed, the awareness for PPCM has risen, simultaneously with new treatments for HF including the use of bromocriptine in this specific setting.

Furthermore, we did not have the full labor data such as fluids administration and rate, as well as the use of labor induction.

Moreover, this study represents a heterogeneous population in Israel, however the validation of the model will not necessarily be the same in other population in different regions of the world.

## Clinical implications

This large-scale study presents the first pre-delivery prediction model for PPCM risk. As some PPCM risk factors are modifiable, our hypothesis is that early understanding and identification of at-risk population (and on the other hand, exclude the surely not-at-risk population) could allow preemptive interventions to reduce disease incidence or change its natural route. Our stratification demonstrates marked heterogeneity in PPCM risk and highlights a small subset of the population with substantially elevated risk who may benefit from targeted preventive interventions.

Therefore, our validated easy to use in-office stratification scoring tool may assist primary care physicians and obstetricians in identifying women at increased risk before pregnancy and supporting tailored monitoring and management, such as closer follow up of designated cardio-obstetric clinic, repeated echocardiography and NT-pro BNP levels, treatment with diuretics and beta blockers accordingly, preliminary discussion about timing and form of labor and hospitalization planning. Such proactive care has the potential to reduce the future incidence or severity of PPCM and improve maternal and fetal outcomes, as well as future family planning and health burden. We also believe that this model will be implacable worldwide and indifferent of insurance status, as all it takes in primary physician and the available score.

## Conclusions

We identified key risk factors associated with PPCM, enabling development of a screening tool for early identification of at-risk women. This model may guide early and intensive follow up and intervention strategies to reduce morbidity and mortality. Our screening protocol tool incorporating these risk factors could enhance clinical vigilance and guide targeted monitoring. Prospective validation of our model is needed to optimize its utility in obstetric and cardiology practice.

## Data Availability

Data will not be available

## Abbreviations

PPCM: Peri partum cardiomyopathy
LV: Left ventricle
LVEF: Left ventricle ejection fraction
SSP: Subsequent pregnancy
HF: Heart failure
CHS: Clalit health services
CVD: Cardiovascular disease
AI: Artificial intelligence
BNP: Beta natriuretic peptide

